# Chest-Pain Descriptors Add Little Diagnostic Value for Coronary Occlusion: A 13,262-Encounter ED Study Using Bayesian Latent-Class Analysis

**DOI:** 10.1101/2025.06.25.25330295

**Authors:** José Nunes de Alencar, Mariana Fuziy Nogueira De Marchi, Kaliana Maria Nascimento Dias de Almeida, Italo Menezes Ferreira, Diandro Marinho Mota, Hugo Ribeiro Ramadan

**Affiliations:** Instituto Dante Pazzanese de Cardiologia. São Paulo. Brazil

**Keywords:** chest pain, acute coronary occlusion, diagnostic accuracy, Bayesian latent-class analysis, emergency department

## Abstract

**Background:** Whether symptom descriptors meaningfully change post-test probability for occlusion myocardial infarction (OMI) in the emergency department (ED) remains uncertain, particularly when the reference standard is imperfect and may miss NSTEMI-OMI without ST elevation.

**Methods:** We retrospectively analyzed 13,262 ED encounters (2021–2025) with chest pain, ECG, and hs-cTnI testing. The index test was a legacy descriptor (type A/B vs C/D). The composite reference was door-to-balloon activation or peak hs-cTnI ≥ 5,000 ng/L, with prespecified sensitivity analyses and a Bayesian latent-class model (BLCA) to correct for reference misclassification. Prognostic value for in-hospital mortality was assessed using patient-clustered multivariable logistic regression with performance and calibration.

**Results:** In frequentist analyses, sensitivity was 0.690 (95% CI 0.657–0.722) and specificity 0.500 (0.491–0.509) (LR+ 1.38, LR− 0.62). BLCA that modeled an imperfect reference yielded a posterior LR+ ≈1.6 and LR− ≈0.43. “Typical” pain did not independently predict mortality after adjustment (adjusted OR 1.13; 95% CI 0.71–1.79).

**Conclusions:** In a large, consecutive ED cohort, chest-pain descriptors offer limited diagnostic leverage for OMI, even after correcting for reference misclassification with BLCA and probing potential missed NSTEMI-OMI.

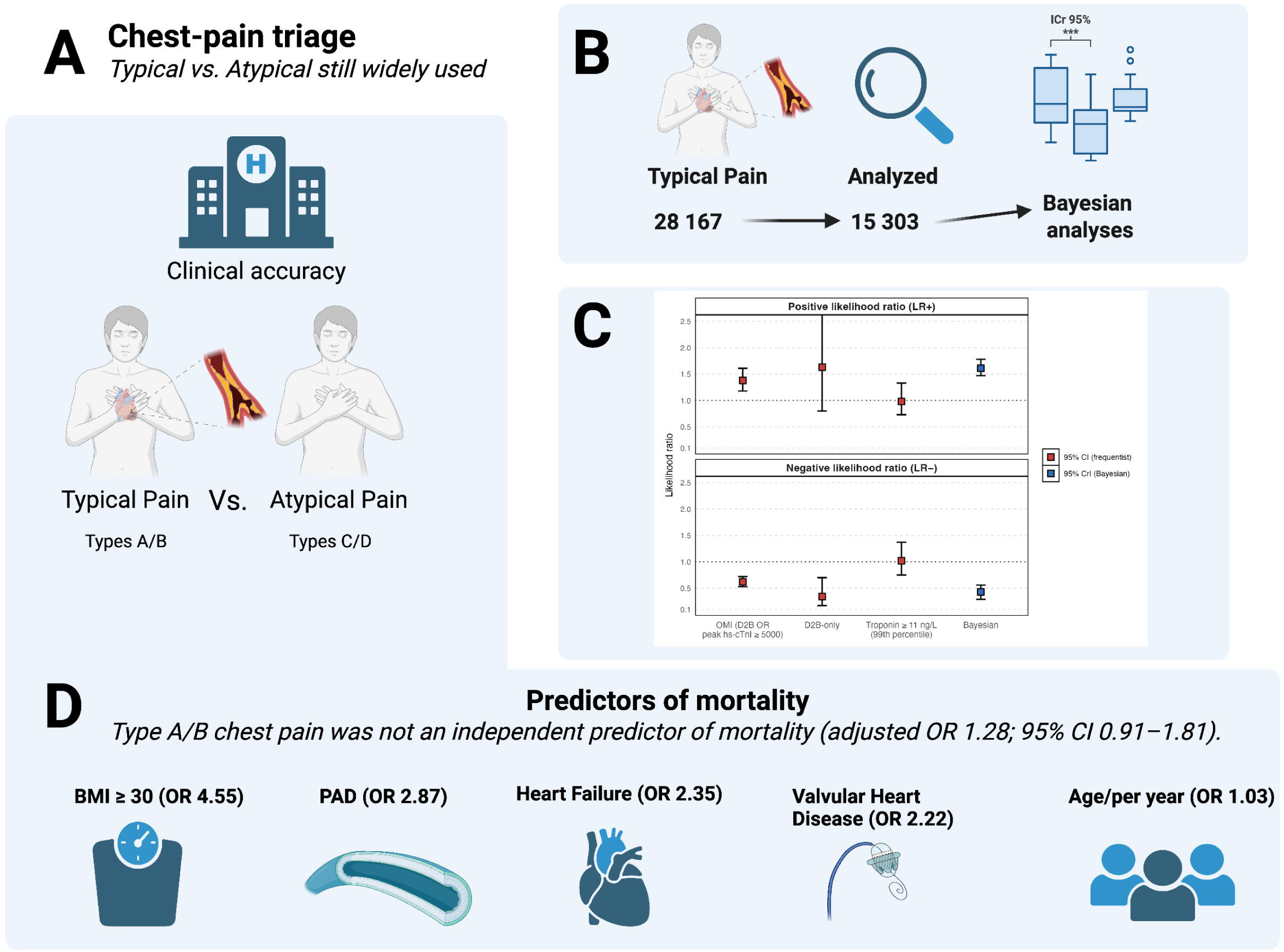

## Introduction

The most recent American guideline on the evaluation of chest pain, the current European guideline on acute coronary syndromes, and the latest American guideline on myocardial infarction each advise against subdividing chest discomfort into the traditional “typical versus atypical”.(1–3) These documents instead recommend describing pain in terms of its pre-test probability of ischemic origin, integrating the full clinical context rather than relying on a rigid semiological grid.

A persistent challenge is that common reference standards for acute coronary occlusion (ACO/OMI)—ST-segment elevation or cath-lab activation—can miss NSTEMI-OMI, and low biochemical cutoffs can misclassify nearly everyone as diseased. This misclassification distorts accuracy estimates for symptom descriptors. We therefore combined a large, consecutive emergency department (ED) dataset with a Bayesian latent-class analysis (BLCA) to account for an imperfect reference standard and to test whether symptom descriptors materially change post-test probability once modern biomarkers are considered.

## Methods

### Study design and setting

We conducted a retrospective observational study in the adult emergency department of a tertiary hospital in São□Paulo, Brazil. All data were gathered between January 2021 and May 2025. The institutional review board approved the protocol with a waiver of individual informed consent because data collection relied exclusively on routinely acquired clinical information. This study follows STARD-BLCM guidelines.(4)

### Patient selection

All consecutive patients aged eighteen years or older who presented with a chief complaint of chest pain were screened. Patients are initially assessed by resident physicians supervised by attending emergency physicians; resident entries are typically verified by attendings. To be eligible, an individual had to undergo a twelve-lead electrocardiogram at presentation and have at least one high-sensitivity cardiac troponin□I (hs-cTn-I) measurement recorded during the index visit. Patients were excluded if either ECG or troponin were not requested.

### Chest-pain classification procedure

At first clinical contact, resident physicians coded the patient’s symptom according to the four⍰tier scheme previously described.(5)□A presentation was labelled “definite angina” (type□A) when the discomfort was retrosternal, precipitated by exertion, radiated to the shoulder, neck, or left arm, and abated within□≈□10□minutes of rest or sublingual nitroglycerin. “Probable angina” (type□B) shared most—but not all—of those features. “Probably non⍰anginal pain” (type□C) denoted atypical chest pain that failed to meet the angina criteria, whereas “definite non⍰anginal pain” (type□D) described discomfort unrelated to physical activity, suggestive of an extracardiac origin and unresponsive to nitrates. Residents were trained in this taxonomy during orientation, and their initial entries in the electronic medical record were usually verified by attending emergency physicians. For analytic purposes we treated the presence of type□A or type□B characteristics (definite or probable angina) as a positive index test (typical angina), whereas type□C or type□D pain was considered negative (atypical angina).

### Definition of acute coronary occlusion

Our primary case definition aimed to capture true-positive occlusions while reducing verification bias. A patient was labelled as having ACO if the emergency team activated the door-to-balloon (D2B) protocol, which obliges emergent percutaneous coronary intervention. Because reliance on D2B alone may overlook occlusions without overt ST-segment elevation, we also classified as cases those whose peak hs-cTn-I reached or exceeded 5000□ng/L. Two pre-specified sensitivity analyses were undertaken: first, repeating all calculations after removing the troponin criterion, and second, redefining cases solely by a peak hs-cTn-I of at least 11□ng/L, the high-risk threshold employed by our laboratory. Controls were patients who did not trigger D2B activation and whose peak troponin remained below 5000□ng/L (or below 11□ng/L in the second sensitivity analysis).

### Biochemical assays

Hs-cTnI was measured using the VITROS platform. Our laboratory reference interval categorizes values below the limit of quantification (LoQ), which is 1.5 ng/L, as very low risk. Values between 1.5 and 10.9 ng/L are considered minor elevations, while values above or equal to 11 ng/L are considered the 99th percentile value.

### Outcomes

We assessed two sets of outcomes. Diagnostic performance—sensitivity, specificity, and positive and negative likelihood ratios—was calculated for the chest-pain test against the reference standard for ACO. Prognostic performance was evaluated by all-cause in-hospital mortality.

### Data handling and statistical analysis

Clinical variables, laboratory values and outcome data were extracted via SQL queries into Microsoft□Excel and analyzed with R version□4.5.0. Categorical variables are presented as counts and proportions; continuous variables as means with standard deviations or medians with inter-quartile ranges, as appropriate. Sensitivity and specificity were accompanied by exact (Clopper-Pearson) 95□percent confidence intervals.(6) Likelihood-ratio confidence intervals were derived with the log-method. Diagnostic metrics were re-computed in prespecified strata defined by sex and by age (<□65□years vs□≥□65□years). For prognostic analysis, we calculated crude odds ratios for death and produced multivariable logistic regression models with patient-clustered robust standard errors, reporting discrimination (AUC with 95% CI), Brier score, and calibration-in-the-large and calibration slope (with 95% CIs). Continuous predictors (e.g., age, log-transformed first hs-cTnI) were scaled per inter-quartile range to aid interpretability. We assessed multicollinearity using variance inflation factors. For each binary comorbidity we constructed a 2 x 2 contingency table and calculated a two-sided Pearson χ² statistic without Yates correction. When any expected frequency fell below five, the χ² approximation was replaced by Fisher’s exact test. Statistical tests were two-sided and a P-value below 0.05 denoted significance.

To account for the recognized misclassification of our composite reference standard, we fitted a four⍰cell multinomial Bayesian latent⍰class model.

- Index test (type□A/B vs□C/D): Se□ ∼ Beta(21, 9) and Sp□ ∼ Beta(15, 16), expressing modest confidence in the accuracy suggested by our frequentist analysis (effective sample size ≈ 30) —an approach recommended by the STARD⍰BLCM extension when reliable internal data exist.
- Composite reference test: Se□ ∼ Beta(6, 5) and Sp□ ∼ Beta(96, 5). For the sensitivity prior (Se□), we adopted a vague Beta(6,□5) distribution, which centers around 0.55 but retains wide uncertainty. This choice reflects the known limitations of our composite case definition. Specifically, the sensitivity of ST-segment elevation (the primary trigger for door-to-balloon activation) for angiographic coronary occlusion has been estimated at just 43.6% in a recent meta-analysis.(7) We complemented this criterion with a peak hs-cTnI ≥□5000□ng/L to capture occlusions that might not manifest ST elevation; however, the diagnostic sensitivity and specificity of that troponin threshold for ACO are not well established in the literature. Given this ambiguity, we intentionally specified a conservative, weakly informative prior that acknowledges substantial uncertainty in the composite reference standard’s ability to detect true occlusions. For specificity (Sp□), we used a more concentrated Beta(96,□5) prior, reflecting high confidence that patients who triggered D2B activation or had a troponin elevation above 5000□ng/L are unlikely to be false positives. This asymmetry in prior strength—vague for sensitivity, concentrated for specificity—was predetermined to align with empirical evidence and clinical expectations in the emergency care setting.
- Prevalence: a Beta (9, 291) prior (mean 0.03, effective sample size = 300) was retained, representing the best external estimate of ACO prevalence in undifferentiated chest-pain populations.
- Sampling and convergence: Three chains were run in rjags (3000 burn⍰in, 9000 retained iterations, thinning□=□3). Convergence was confirmed by Gelman–Rubin R⍰hat ≤□1.002, visual trace⍰plot stability, effective sample sizes□>□3700 for every target, and time⍰series Monte⍰Carlo SE□≤□0.0033. Posterior medians and equa⍰ltail 95% credible intervals (CrI) are reported.
- Sensitivity to conditional dependence: Because the index test and the composite reference share semiological information, we performed a secondary Bayesian run assuming a modest positive conditional correlation (ρ□=□0.20) between their latent errors, implemented with the Sarmanov method. All other priors and sampling settings were left unchanged.

### Sensitivity analysis

To probe the robustness of all frequentist estimates, we re-ran the complete diagnostic work-up under three prespecified scenarios: (1) **reference-standard restriction**, limiting the gold standard to door-to-balloon (D2B) activation alone; (2) **biochemical benchmark**, defining cases exclusively by a peak high-sensitivity troponin ≥ 11 ng/L; and (3) **alternative index-test definition**, narrowing a “positive” index test to **type A** chest pain only (types **B/C/D** pooled as negative). In an additional prespecified robustness check, we performed a **threshold sweep** for the composite reference standard (D2B **or** peak troponin) across cutoffs of 1,000/2,000/5,000/10,000 ng/L and recomputed accuracy metrics at each threshold.

## Results

Among 28167 emergency-department visits in which troponin and an EKG were requested in 2021–2025, 395 had < 18 years, 1184 were transferred from another hospital, and 9978 were excluded due to no reference to types A, B, C or D or every box was ticked “no”. The remaining 13262 encounters, from 11258 unique patients, constituted the analytic cohort (Figure 1).

**Figure 1.**
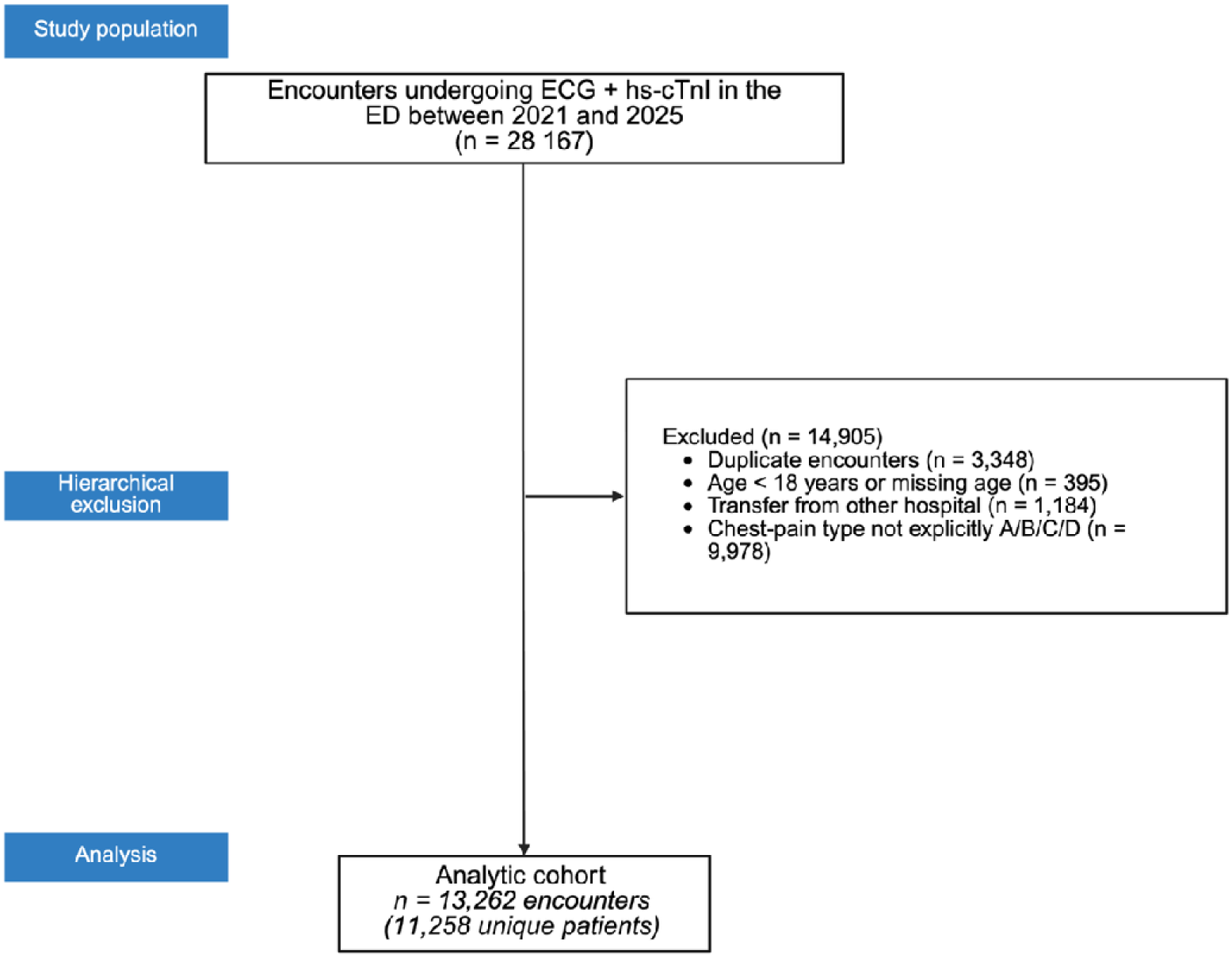
Flow of patients through the study.

The median age of these patients was 62 years (inter-quartile range 52–71). Men accounted for 53.6 % of the population and women for 46.4 %. Additional baseline characteristics are summarized in Table 1.

**TABLE 1.** Baseline prevalence of selected comorbidities among ACO “cases” (door-to-balloon activation and/or peak hs-cTnI ≥ 5000 ng L⁻^¹^) versus “controls” (no ACO). Categorical variables are presented as n (%). p-values derive from a two-sided Pearson χ² test; Fisher’s exact test was used when any expected cell count was < 5. Missing values (e.g., BMI) were excluded from each comparison.

| Variable | Cases n (%) | Controls n (%) | p-value |
| --- | --- | --- | --- |
| Age, median (IQR), y | 65 [58–73] | 62 [52–70] | <0.0001 |
| Female sex | 272 (33.4%) | 5883 (47.3%) | <0.0001 |
| hs-cTnl, first (ng/L), median (IQR) | 6342.5 [4440.8–9260.5] | 139.0 [15.0–522.0] | <0.0001 |
| hs-cTnl, peak (ng/L), median (IQR) | 7851.5 [6168.5–12450.0] | 163.5 [15.0–579.0] | <0.0001 |
| Door-to-balloon activation | 54 (6.6%) | 0 (0.0%) | <0.0001 |
| BMI ≥ 30 | 118/451 (26.2%) | 328/1127 (29.1%) | 0.2413 |
| Smoking | 143/564 (25.4%) | 1369/8693 (15.7%) | <0.0001 |
| Stroke | 51 (6.3%) | 702 (5.6%) | 0.4561 |
| Hypertension | 655 (80.5%) | 8945 (71.9%) | <0.0001 |
| Diabetes Mellitus | 238/712 (33.4%) | 3278/11624 (28.2%) | 0.0027 |
| Dyslipidemia | 486 (59.7%) | 6686 (53.7%) | 0.0009 |
| Peripheral Arterial Disease | 32 (3.9%) | 255 (2.0%) | 0.0003 |
| Valvular Heart Disease | 38 (4.7%) | 804 (6.5%) | 0.0422 |
| Heart Failure | 170 (20.9%) | 2140 (17.2%) | 0.0072 |
| Coronary Artery Disease | 431 (52.9%) | 4761 (38.2%) | <0.0001 |
| Prior PCI | 142 (17.4%) | 1563 (12.6%) | <0.0001 |
| Prior CABG | 86 (10.6%) | 803 (6.5%) | <0.0001 |
| Chronic Kidney Disease | 91 (11.2%) | 755 (6.1%) | <0.0001 |
| Chagas Disease | 10 (1.2%) | 157 (1.3%) | 0.9353 |

### Diagnostic performance in frequentist estimates

When the composite reference standard was applied, the 2 × 2 table consisted of 562 true positives, 252 false negatives (814 cases), 6 226 false positives, and 6 222 true negatives (12 448 controls). From these figures, the “type A or B” (typical) designation yielded a sensitivity of 0.690 (95 % CI 0.657–0.722) and a specificity of 0.500 (95 % CI 0.491–0.509). The positive likelihood ratio (LR+) was 1.38 (95 % CI 1.18–1.61) and the negative likelihood ratio (LR−) 0.62 (95 % CI 0.53–0.72).

Sex stratification produced broadly comparable figures. In women, sensitivity was 0.680 (95 % CI 0.621–0.735) and specificity 0.542 (95 % CI 0.529–0.555); LR+ 1.49 (95 % CI 1.15–1.93) and LR− 0.59 (95 % CI 0.46–0.77). In men, sensitivity reached 0.696 (95 % CI 0.655–0.734) and specificity 0.462 (95 % CI 0.450–0.474); LR+ 1.29 (95 % CI 1.07–1.56) and LR− 0.66 (95 % CI 0.55–0.80).

Accuracy did not change with age. Participants younger than sixty-five years showed a sensitivity of 0.687 and specificity of 0.513; LR+ 1.41 (95 % CI 1.13–1.76) and LR− 0.61 (95 % CI 0.49–0.76). For those sixty-five years or older, sensitivity was 0.694 and specificity 0.482; LR+ 1.34 (95 % CI 1.08–1.66) and LR− 0.63 (95 % CI 0.51–0.79).

### Sensitivity analysis

When the reference standard was restricted to D2B activation alone, the 2 × 2 table contained 45 true positives, 9 false negatives, 6743 false positives and 6465 true negatives. Sensitivity was 0.833 (95 % CI 0.707–0.921) while specificity was 0.489 (95 % CI 0.481–0.498). LR+ was 1.63 (95 % CI 0.80–3.34) and LR− 0.34 (95 % CI 0.17–0.70).

We re-specified the index test to treat type A pain alone as “positive,” lumping types B, C, and D together as “negative.” Under the composite reference standard this definition achieved a sensitivity of 0.316 (95% CI 0.284–0.349) and a markedly higher specificity of 0.859 (95% CI 0.852–0.865). The resulting LR+ rose to 2.23 (95% CI 1.91–2.61) and LR− was 0.80 (95% CI 0.68–0.93).

Using peak hs-cTnI ≥ 11 ng/L as the disease definition, the 2×2 table yielded 6,698 true positives, 6,391 false negatives, 90 false positives, and 83 true negatives. Sensitivity was 0.512 (95% CI 0.503–0.520) and specificity 0.480 (95% CI 0.403–0.557). The likelihood ratios were uninformative: LR+ 0.98 (95% CI 0.73–1.33) and LR− 1.02 (95% CI 0.75–1.37). Notably, the prevalence was 98.7% of analyzed encounters, indicating that this low biochemical threshold classifies nearly the entire cohort as “diseased,” which in turn drives the LRs toward 1 and limits diagnostic usefulness. Finally, a threshold sweep across 1,000/2,000/5,000/10,000 ng/L showed the same qualitative pattern, with LR+ in the ∼1.35–1.49 range and LR− in the ∼0.49–0.69 range (e.g., at 10,000 ng/L: sensitivity 0.756 [95% CI 0.701–0.806], specificity 0.493 [95% CI 0.485–0.502], LR+ 1.49 [95% CI 1.13–1.97], LR− 0.49 [95% CI 0.37–0.65]).

### Diagnostic performance in Bayesian estimates

Bayesian latent-class analysis that incorporated weighted priors for the imperfect reference standard produced a posterior median sensitivity of 0.778 (95 % CrI 0.714–0.849) and a posterior median specificity of 0.517 (95 % CrI 0.505–0.531) for the type A/B designation. The corresponding median LR+ was 1.609 (95 % CrI 1.466–1.779), whereas the median LR− was 0.430 (95 % CrI 0.289–0.555). All Gelman–Rubin R-hat statistics were ≤ 1.004, effective sample sizes were ≥ ∼780 for all targets, and chains mixed well. Results are summarized in Figure 2.

**Figure 2.**
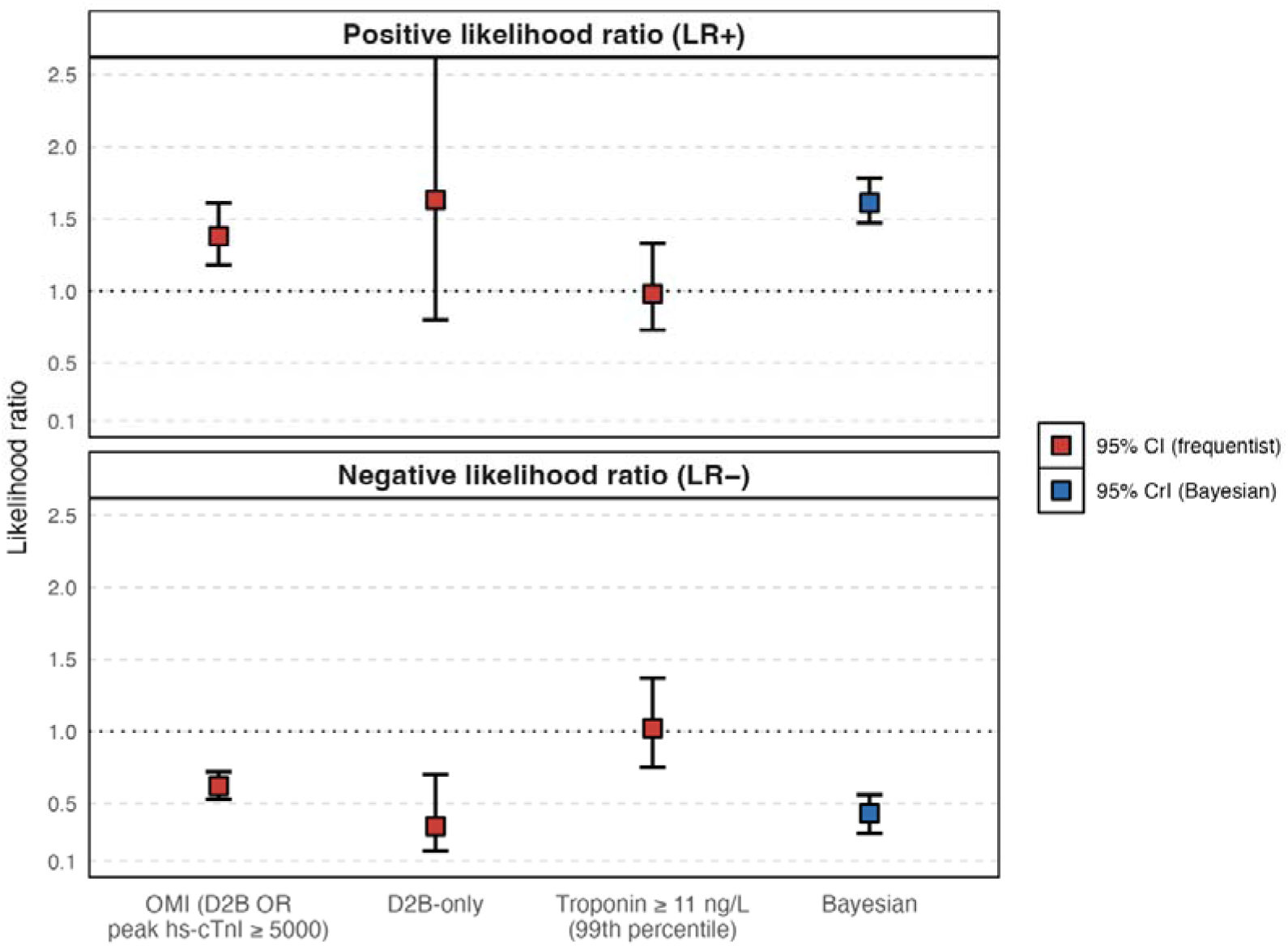
Positive and negative likelihood ratios for chest-pain type A/B (typical) under different reference definitions. Red markers and vertical lines represent the frequentist point estimates and 95% confidence intervals; blue markers and vertical lines represent Bayesian posterior medians and 95% credible intervals. From left to right: (1) door-to-balloon (D2B) activation and/or peak hs-cTnI ≥ 5000 ng L⁻^¹^, (2) D2B activation only, (3) peak troponin ≥ 99th percentile, and (4) Bayesian latent-class model (ρ = 0). The horizontal dashed line at LR⁺ = 1.0 indicates no change in post-test probability.

Regarding conditional dependence, when a correlation of ρ = 0.20 was imposed between the errors of the index and reference tests, the posterior median sensitivity was 0.782 (95 % CrI 0.716–0.852) and specificity 0.515 (95 % CrI 0.504–0.529); LR+ 1.613 (95 % CrI 1.463–1.782) and LR− 0.423 (95 % CrI 0.284–0.555). Convergence remained excellent. These figures are essentially superimposable on the base-case model, indicating that modest conditional dependence does not materially alter the diagnostic verdict.

### Prognostic performance

There were 150 in-hospital deaths among the analytic cohort. In univariate models, chest-pain classified as typical (versus atypical) was associated with higher mortality (OR 1.77; 95% CI 1.17–2.74). However, after adjusting for covariates in a multivariable model with patient-clustered standard errors, the association between type A/B pain and death was attenuated and no longer statistically significant (adjusted OR 1.13; 95% CI 0.71–1.79). A BMI ≥ 30 kg/m² remained independently associated with in-hospital mortality (adjusted OR 1.99; 95% CI 1.02–3.90). Heart failure (adjusted OR 1.75; 95% CI 1.10–2.76) and valvular heart disease (adjusted OR 2.00; 95% CI 1.04–3.85) were also independently associated with death, whereas peripheral artery disease showed a non-significant trend (adjusted OR 1.98; 95% CI 0.91–4.34). (Table 2).

**TABLE 2.** Univariate and multivariable logistic regression analyses evaluating predictors of in-hospital mortality. (Age and first troponin are scaled per IQR for interpretability; first troponin uses log10.)

| Variable | Univariate<br>OR | Univariate<br>95% CI | Multivariable<br>OR | Multivariable<br>95% CI |
| --- | --- | --- | --- | --- |
| Chest pain A/B vs.<br>C/D | 1.77 | 1.17 – 2.74 | 1.13 | 0.71 – 1.79 |
| IMC ≥ 30 | 6.08 | 3.40 – 10.2 | 1.99 | 1.02 – 3.90 |
| Peripheral Artery<br>Disease | 3.70 | 1.54 – 7.51 | 1.98 | 0.91 – 4.34 |
| Heart Failure | 2.84 | 1.85 – 4.30 | 1.75 | 1.10 – 2.76 |
| Chagas Disease | 3.55 | 1.07 – 8.61 | 2.57 | 0.90 – 7.36 |
| Chronic Kidney<br>Disease | 3.53 | 2.04 – 5.79 | 1.30 | 0.71 – 2.37 |
| Valvular Heart<br>Disease | 2.18 | 1.12 – 3.85 | 2.00 | 1.04 – 3.85 |
| Hypertension | 3.22 | 1.76 – 6.63 | 1.48 | 0.69 – 3.19 |
| Stroke | 1.77 | 0.82 – 3.34 | – | – |
| Dyslipidemia | 2.36 | 1.51 – 3.80 | 1.19 | 0.69 – 2.06 |
| Diabetes Mellitus | 1.89 | 1.24 – 2.85 | 1.29 | 0.83 – 2.02 |
| Coronary Artery<br>Disease | 1.93 | 1.29 – 2.92 | 1.03 | 0.67 – 1.59 |
| Age (per IQR) | 2.50 | 1.82 – 3.45 | 1.70 | 1.17 – 2.48 |
| First troponin<br>(log10), per IQR | 16.3 | 10.8 – 24.8 | 10.3 | 4.85 – 22.0 |
| Smoking | 1.48 | 0.82 – 2.50 | – | – |
| Sex (Male) | 1.22 | 0.81 – 1.86 | 0.87 | 0.56 – 1.36 |

**TABLE 3.**
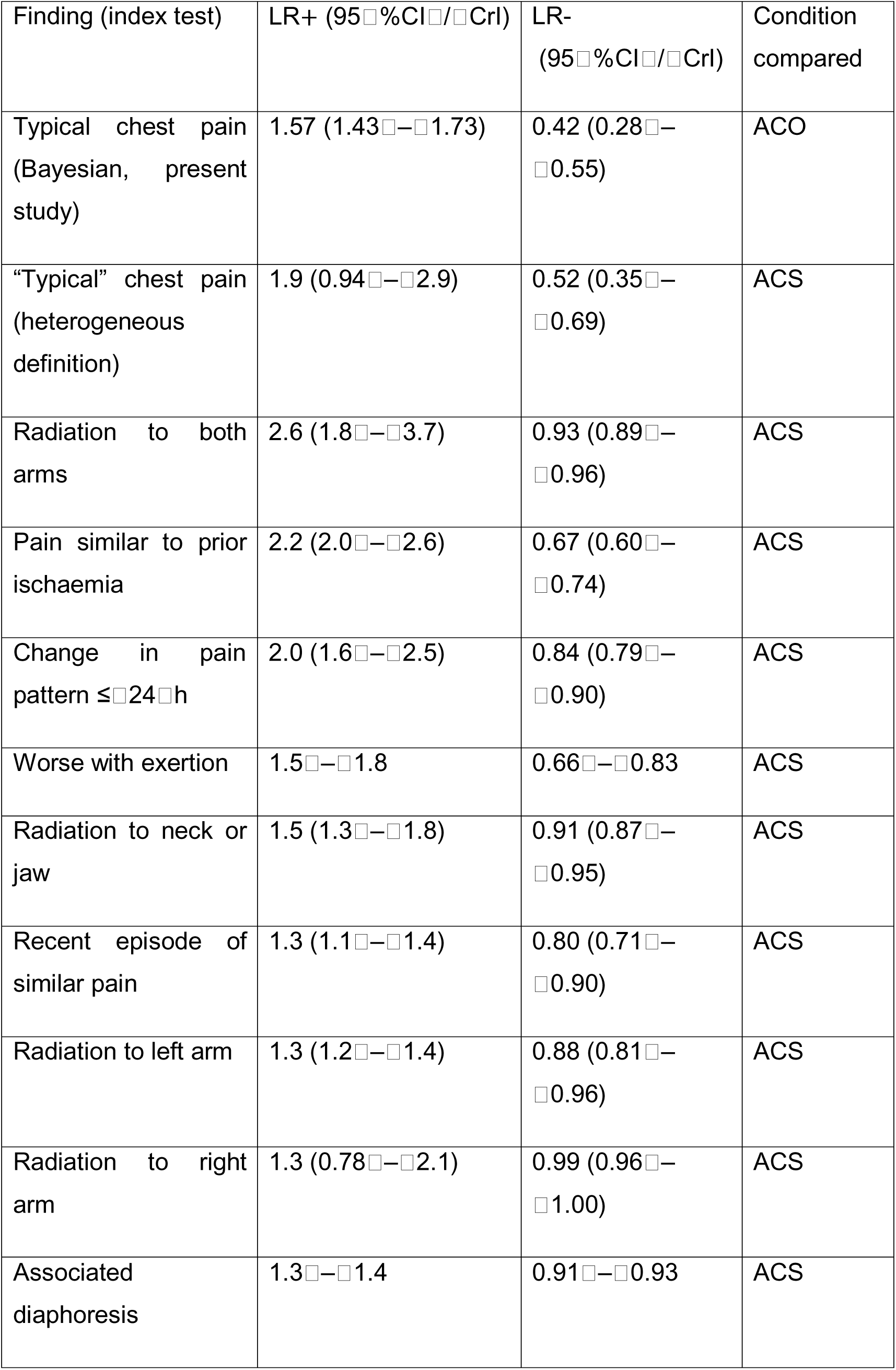
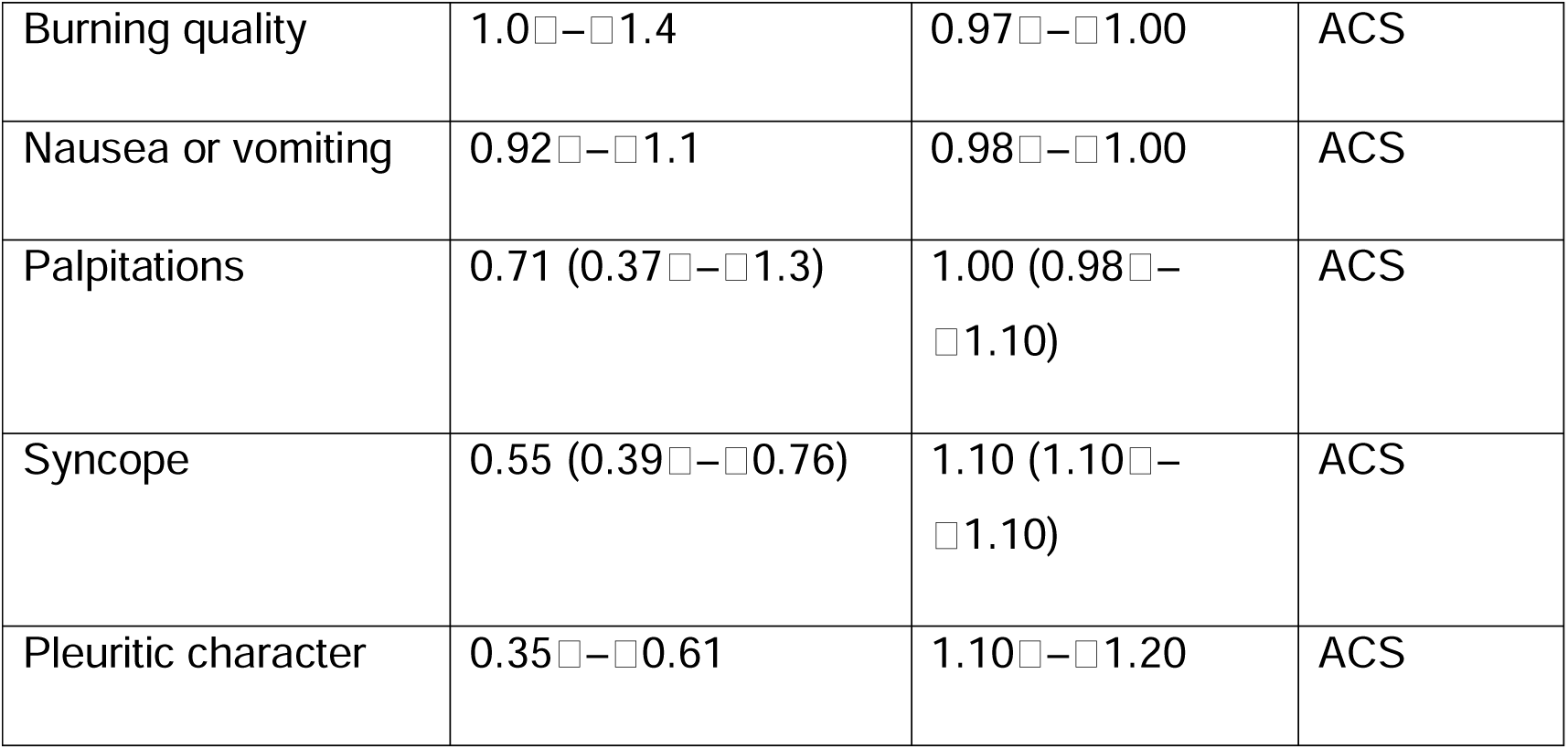
Selected chest⍰pain descriptors (from Fanaroff’s meta-analysis)(13) and their diagnostic value. Abbreviations: LR+□=□positive likelihood ratio; LR-□=□negative likelihood ratio; ACS□=□acute coronary syndrome (definitions varied across source studies); ACO□=□acute coronary occlusion (door⍰to⍰balloon activation or peak hs⍰cTnI□≥□5000□ng/L in the present study). Ranges without parentheses indicate the span of individua⍰lstudy estimates when a pooled CI was not reported.

| Finding (index test) | LR+ (95%CI/CrI) | LR- (95%CI/CrI) | Condition compared |
| --- | --- | --- | --- |
| Typical chest pain (Bayesian, present study) | 1.57 (1.43–1.73) | 0.42 (0.28–0.55) | ACO |
| “Typical” chest pain (heterogeneous definition) | 1.9 (0.94–2.9) | 0.52 (0.35–0.69) | ACS |
| Radiation to both arms | 2.6 (1.8–3.7) | 0.93 (0.89–0.96) | ACS |
| Pain similar to prior ischaemia | 2.2 (2.0–2.6) | 0.67 (0.60–0.74) | ACS |
| Change in pain pattern $\leq 24$ h | 2.0 (1.6–2.5) | 0.84 (0.79–0.90) | ACS |
| Worse with exertion | 1.5–1.8 | 0.66–0.83 | ACS |
| Radiation to neck or jaw | 1.5 (1.3–1.8) | 0.91 (0.87–0.95) | ACS |
| Recent episode of similar pain | 1.3 (1.1–1.4) | 0.80 (0.71–0.90) | ACS |
| Radiation to left arm | 1.3 (1.2–1.4) | 0.88 (0.81–0.96) | ACS |
| Radiation to right arm | 1.3 (0.78–2.1) | 0.99 (0.96–1.00) | ACS |
| Associated diaphoresis | 1.3–1.4 | 0.91–0.93 | ACS |
| Burning quality | 1.0□–□1.4 | 0.97□–□1.00 | ACS |
| Nausea or vomiting | 0.92□–□1.1 | 0.98□–□1.00 | ACS |
| Palpitations | 0.71 (0.37□–□1.3) | 1.00 (0.98□–<br>□1.10) | ACS |
| Syncope | 0.55 (0.39□–□0.76) | 1.10 (1.10□–<br>□1.10) | ACS |
| Pleuritic character | 0.35□–□0.61 | 1.10□–□1.20 | ACS |

Age remained independently predictive of death—per-IQR increase in age was associated with a 70% increase in odds (adjusted OR 1.70; 95% CI 1.17–2.48). In contrast, first troponin (log10), per IQR showed a strong independent association with mortality (adjusted OR 10.3; 95% CI 4.85–22.0), while sex and smoking were not independently associated in the fully adjusted model. The final model demonstrated excellent discrimination (AUC 0.902; 95% CI 0.875–0.929) and low overall error (Brier score 0.0068). Calibration-in-the-large was essentially null (intercept −0.000; 95% CI −0.209 to 0.209) and the calibration slope was near-ideal (1.000; 95% CI 0.860–1.140). Multicollinearity was not concerning (all VIFs ≤ 1.91).

## Discussion

The latest U.S. chest⍰pain guideline explicitly discourages a dichotomous “typical versus atypical” mindset and urges clinicians to weigh individual semiological elements and contextual probability.(1) Our data show that the global label “typical” conveys very limited incremental information when modern biochemical and angiographic standards are applied.

Two features distinguish this study from prior reports. First, scale—a consecutive, real-world ED cohort large enough to estimate precision credibly. Second, method—we used BLCA to address the known imperfection of ST-elevation/cath activation and of low troponin cutoffs as ‘truth.’ Together, these design choices reduce both work-up and incorporation biases and directly tackle the problem of missed NSTEMI-OMI.

Bassan and colleagues(8) reported a sensitivity of 94% and a specificity of 52% for the A/B schema for typical pain in 2000, findings widely quoted in Brazilian textbooks and triage protocols.□However, their case definition required an elevated CK⍰MB, which was not measured in patients assigned to type□D, thereby locking the semiology into both incorporation and work⍰up bias.□By contrast, we defined acute coronary occlusion as either a D2B activation with angiographic evidence of an occluded vessel or a peak high⍰sensitivity troponin□I ≥□5000□ng/L.(9)□This dual criterion recognizes that ST⍰segment–elevation is an imperfect surrogate for occlusion and partially mitigates the risk of missing non⍰ST⍰elevation occlusions that never reach the catheterization laboratory.(10) In prespecified sensitivity analyses, dropping the troponin criterion (D2B-only) produced sensitivity 0.833 (95% CI 0.707–0.921) and specificity 0.489 (95% CI 0.481–0.498), while redefining “positive” pain as type A alone increased specificity to 0.859 (95% CI 0.852–0.865) at the cost of sensitivity 0.316 (95% CI 0.284–0.349). A purely biochemical benchmark (peak hs-cTnI ≥ 11 ng/L) classified ∼98.7% of encounters as “diseased,” yielding sensitivity 0.512 (95% CI 0.503–0.520), specificity 0.480 (95% CI 0.403–0.557), and likelihood ratios ∼1, which are clinically uninformative. Finally, a threshold sweep across 1,000/2,000/5,000/10,000 ng/L showed LR+ in the ∼1.35–1.49 range and LR− in the ∼0.49–0.69 range (e.g., at 10,000 ng/L: sensitivity 0.756 [0.701–0.806], specificity 0.493 [0.485–0.502], LR+ 1.49 [1.13–1.97], LR− 0.49 [0.37–0.65]). Together, these experiments reinforce that the A/B construct adds, at best, modest diagnostic leverage. Although our index test used a legacy, locally implemented four-tier symptom taxonomy, the clinical message generalizes: global pain labels add minimal diagnostic value once ECG and hs-cTnI are available.

A more recent registry study of unstable angina found that type□A pain predicted the decision to pursue invasive stratification.(11)□That observation likely represents a form of collinearity bias: the semiology helps drive the decision to catheterize, and catheterization then becomes the outcome under study.□Our design, anchored in objective biochemical thresholds and procedural activations avoids this circularity.

Notably, the American guideline cites a likelihood ratio of 4.26 for pain radiating to both arms—one isolated descriptor— whereas our primary A/B definition produced LR+ 1.38 and LR− 0.62 under the composite standard, and even our stricter “type A only” definition reached only LR+ 2.23 with LR− 0.80. Thus, although “type A only” improves rule-in performance relative to A/B, it remains far below the >10 LR+ benchmark for confident rule-in, and our best LR− values (as low as 0.34 under D2B-only) remain well above the <0.1 threshold for confident rule-out..(12)□TableC3 juxtaposes our key likelihood ratios with those thresholds.(13)

A key strength of our work is the application of a Bayesian latent⍰class model to account for misclassification within the composite reference standard. This approach allowed the data to “speak for itself” by integrating prior knowledge— specifically, modest confidence in the index test’s accuracy combined with more certain expectations regarding the specificity of the composite reference test— into a cohesive probabilistic framework. After accounting for misclassification and adopting a low but plausible prior prevalence (∼3%), the posterior medians were LR+ 1.61 (95% CrI 1.47–1.78) and LR− 0.43 (95% CrI 0.29–0.56). Those figures closely mirror the best⍰available aggregated evidence: Fanaroff’s landmark meta⍰analysis of six heterogeneous studies (≈□14000 patients, I²□=□98□%) reported an overall LR+ of□1.9□(95□%□CI□0.94–2.9) and an LR-of□0.52□(95□%□CI□0.35–0.69).(13)□ Assuming modest positive conditional dependence (ρ = 0.20) between index and reference tests left the posterior essentially unchanged. The wide heterogeneity in that review stemmed mostly from inconsistent definitions of “typical” pain—an observation that links with our finding that type⍰based labels encode far more clinician gestalt than textbook criteria.□Indeed, the modest Bayesian LR-in our data likely reflects the tacit synthesis physicians apply when designating pain as type□C or□D—subtle vocal inflections, patient behavior, risk⍰factor burden and on⍰the⍰spot ECG interpretation—much as Bassan’s original four⍰tier construct captured nuances no rigid checklist could codify.(8) Our Bayesian model, by letting the data rather than a single “truth” table guide the posterior, may therefore be tapping into that clinical intuition. Even so, whatever tacit knowledge is wrapped into the A/B label, it does not propel LR+ anywhere near the >□10 threshold needed for a confident rule⍰in, and its potential rule⍰out value remains unacceptably fragile.

### Prognostic perspective

In univariate analysis, typical (A/B) pain was associated with higher in-hospital mortality (OR 1.77; 95% CI 1.17–2.74). However, in a multivariable model with patient-clustered standard errors, the association disappeared (adjusted OR 1.13; 95% CI 0.71–1.79). BMI ≥ 30 kg/m² (adjusted OR 1.99; 95% CI 1.02–3.90), heart failure (adjusted OR 1.75; 95% CI 1.10–2.76), and valvular heart disease (adjusted OR 2.00; 95% CI 1.04–3.85) remained independently associated with death; peripheral artery disease showed a non-significant trend (adjusted OR 1.98; 95% CI 0.91–4.34). Per-IQR age was associated with a 70% increase in odds (adjusted OR 1.70; 95% CI 1.17–2.48), and per-IQR log10 first hs-cTnI showed a strong independent association (adjusted OR 10.3; 95% CI 4.85–22.0).

### Strengths and limitations

The main limitation of the present work is the very small proportion of D2B activations in the analytic cohort—54 of 13 262 encounters (0.41%)—which limits power for D2B-only analyses and widens precision around estimates. A second limitation stems from case adjudication: ideally, an OMI/ACO diagnosis is established prospectively from high-fidelity data—serial high-sensitivity troponin curves (“delta”), contemporaneous ECG interpretation, detailed coronary angiography (including TIMI flow), and adjunct imaging. In a retrospective database many of these elements are incomplete or unavailable, including systematic catheterization results, compelling a pragmatic composite reference standard that combined door-to-balloon activation (procedural trigger for emergent PCI) with a peak hs-cTnI ≥ 5 000 ng/L. We adopted D2B because it directly operationalizes an ED decision to revascularize and is captured reliably in the EHR, whereas angiographic occlusion metrics were inconsistently recorded. Although this strategy mitigates classic work-up bias, it can admit false positives (e.g., non-occlusive etiologies with very high troponin) and almost certainly misses some false-negative occlusions (e.g., NSTEMI-OMI without D2B activation).

The choice of 5 000 ng/L for peak hs-cTnI was not intended to mirror the Universal Definition but to enrich for large infarction/likely occlusion. We explicitly stress-tested this threshold: (i) the distribution of the first hs-cTnI— independent of the reference—showed controls rarely approach 5 000 ng/L while cases frequently exceed it, supporting its use as a rule-in enrichment; and (ii) a prespecified threshold sweep (1 000/2 000/5 000/10 000 ng/L) showed that our central conclusion—only modest LR+ and LR− for the A/B label—was qualitatively unchanged across cutoffs. Conversely, using the laboratory 99th-percentile benchmark (hs-cTnI ≥ 11 ng/L) rendered 98.7% of analyzed encounters “diseased,” driving LR estimates toward 1 and underscoring why a low biochemical threshold is unsuitable as an occlusion reference in this context.

Missing data are substantial: 44% of otherwise eligible visits lacked usable A–D coding (no chest-pain type recorded or every box ticked “no”). This may introduce bias if documentation was not missing at random (e.g., sicker or busier encounters less likely to be coded). While this reflects real-world documentation habits and could enhance external validity, it reduces internal completeness.

Symptom classification was performed by resident physicians and usually verified by attendings, but we did not measure inter-rater reliability, leaving room for misclassification of the index test. The ED staffing model (resident–attending) and training may differ from other systems, tempering generalizability. We also collapsed the ordinal A–D scale to a binary “A/B vs C/D” construct to reflect how the terminology is applied operationally at triage; nevertheless, this sacrifices ordinal information. To address the reviewer’s concern, we ran a sensitivity analysis designating “type A only” as positive, which increased specificity to 0.859 at the expense of sensitivity 0.316, but did not meaningfully elevate LR+ to a clinically decisive range.

The study period (2021–2025) spans the COVID-19 era, during which ED volumes, testing patterns, and causes of troponin elevation may have shifted; we could not fully adjust for these secular changes. Single-center design (tertiary hospital, São Paulo) further limits generalizability. We did not adjudicate a subsample of charts to the 4th Universal Definition of MI, which could have better informed priors for our Bayesian model; instead, we specified conservative, transparent priors and demonstrated that posterior inferences were stable even under modest conditional dependence. Finally, as with any retrospective EHR study, data-capture and coding errors are possible despite standardized SQL extraction and audit checks.

These limitations must be weighed against several strengths. First, the cohort is consecutive and unselected with explicit, pre-registered filtering steps—a close approximation of everyday ED practice. Second, incorporation bias was minimized (the pain label never entered the case definition), and work-up bias was mitigated by including a biochemical criterion to capture occlusions not taken for emergent PCI. Third, we executed a comprehensive sensitivity program (D2B-only, hs-cTnI ≥ 11 ng/L, “type A-only” index, and multi-threshold sweeps) and a Bayesian latent-class analysis to account for reference misclassification; all approaches converged on modest LR+ and LR− for the A/B schema. Fourth, prognostic modeling incorporated patient-clustered robust SEs, per-IQR scaling of continuous predictors, and full performance reporting (AUC, Brier, calibration), with no concerning multicollinearity. Together, these features reinforce the practical message: the global “typical vs atypical” label adds limited diagnostic leverage for acute coronary occlusion and does not independently predict in-hospital mortality once core covariates are considered.

## Conclusions

In a consecutive cohort of more than fifteen thousand emergency⍰department presentations, the traditional typical versus atypical chest⍰pain designation identified acute coronary occlusion with a sensitivity of roughly 68% but a specificity of 48%, translating to likelihood ratios (LR+C≈C1.3;CLR-C≈C0.6) that scarcely altered pre⍰test probability.CThese metrics remained weak across sex and age strata and under three alternative case definitions.

A Bayesian latent⍰class analysis that explicitly corrected for misclassification in the reference standard reached the same clinical result: the posterior LR+ centered at□1.5 and the posterior LR-at□0.4. Taken together with current guideline advice and earlier methodological critiques, these data show that typical versus atypical classification confers negligible diagnostic or prognostic value in modern practice. Emergency departments should retire global pain labels and instead anchor triage on granular symptom descriptors, rapid high⍰sensitivity biomarkers and validated risk tools.

## Author approval

All authors have seen and approved the manuscript.

## Data availability statement

Due to restrictions imposed by the institutional review board (IRB) related to patient confidentiality and data anonymization procedures, the datasets analyzed in this study are not publicly available.

## Ethics approval

IRB number 76670223.4.0000.5462

## Patient consent

N/A

## Clinical trial registration

N/A

## Permission to reproduce material from other sources

N/A

## Author Contributions

- **Conceptualization:** José Nunes de Alencar Neto (JNdA), Mariana F. N. De Marchi (MFNDM)
- Project Administration: JNdA, MFNDM
- Methodology & Formal Analysis: JNdA
- **Investigation / Data Curation:** JNdA, Kaliana M. N. Dias de Almeida (KMDdA)
- Writing –Original Draft: JNdA
- **Writing – Review & Editing:** JNdA, MFNDM, KMDdA, Italo Menezes Ferreira (IMF), Diandro Marinho Mota (DMM), Hugo Ribeiro Ramadan (HRR)
- Guarantor of the Article: JNdA accepts full responsibility for the integrity of the study and the manuscript.

All authors had full access to the data, substantially contributed to manuscript revision, and approved the final version.

